# Aerosol emission from the respiratory tract: an analysis of relative risks from oxygen delivery systems

**DOI:** 10.1101/2021.01.29.21250552

**Authors:** F Hamilton, F Gregson, D Arnold, S Sheikh, K Ward, J Brown, E Moran, C White, A Morley, AERATOR group, B Bzdek, J Reid, N Maskell, JW Dodd

**Affiliations:** Infection Sciences, North Bristol NHS Trust, BS10 5NB; MRC Integrative Epidemiology Unit, University of Bristol, BS8 2HW; Bristol Aerosol Research Centre, School of Chemistry, University of Bristol, BS8 1TS; Academic Respiratory Unit, University of Bristol, BS10 5NB; Physiotherapy Department, North Bristol NHS Trust, BS10 5NB; Intensive Care Unit, North Bristol NHS Trust, BS10 5NB; Research and Innovation, North Bristol NHS Trust, BS10 5NB

## Abstract

**Background:** Risk of aerosolisation of SARS-CoV-2 directly informs organisation of acute healthcare and PPE guidance. Continuous positive airways pressure (CPAP) and high-flow nasal oxygen (HFNO) are widely used modes of oxygen delivery and respiratory support for patients with severe COVID-19, with both considered as high risk aerosol generating procedures. However, there are limited high quality experimental data characterising aerosolisation during oxygen delivery and respiratory support.

**Methods:** Healthy volunteers were recruited to breathe, speak, and cough in ultra-clean, laminar flow theatres followed by using oxygen and respiratory support systems. Aerosol emission was measured using two discrete methodologies, simultaneously. Hospitalised patients with COVID-19 were also recruited and had aerosol emissions measured during breathing, speaking, and coughing.

**Findings:** In healthy volunteers (n = 25 subjects; 531 measures), CPAP (with exhalation port filter) produced less aerosols than breathing, speaking and coughing (even with large >50L/m facemask leaks). HFNO did emit aerosols, but the majority of these particles were generated from the HFNO machine, not the patient. HFNO-generated particles were small (<1μm), passing from the machine through the patient and to the detector without coalescence with respiratory aerosol, thereby unlikely to carry viral particles. Coughing was associated with the highest aerosol emissions with a peak concentration at least 10 times greater the mean concentration generated from speaking or breathing. Hospitalised patients with COVID-19 (n = 8 subjects; 56 measures) had similar size distributions to healthy volunteers.

**Interpretation:** In healthy volunteers, CPAP is associated with less aerosol emission than breathing, speaking or coughing. Aerosol emission from the respiratory tract does not appear to be increased by HFNO. Although direct comparisons are complex, cough appears to generate significant aerosols in a size range compatible with airborne transmission of SARS-CoV-2. As a consequence, the risk of SARS-CoV-2 aerosolisation is likely to be high in all areas where patients with Covid-19 are coughing. Guidance on personal protective equipment policy should reflect these updated risks.

**Funding:** NIHR-UKRI Rapid COVID call (COV003), Wellcome Trust GW4-CAT Doctoral Training Scheme (FH), MRC CARP Fellowship(JD, MR/T005114/1). Natural Environment Research Council grant (BB, NE/P018459/1)

**Research in context:** *Evidence before this study:* PubMed was searched from inception until 10/1/21 using the terms ‘aerosol’, and variations of ‘non-invasive positive pressure ventilation’ and ‘high-flow nasal oxygen therapy’. Studies were included if they measured aerosol generated from volunteers or patients receiving non-invasive positive pressure ventilation (NIV) or high flow nasal oxygen therapy (HFNO), or provided experimental evidence on a simulated human setting. One study was identified (Gaeckle et al, 2020) which measured aerosol emission with one methodology (APS) but was limited by high background concentration of aerosol and a low number of participants (n = 10).

*Added value of this study:* This study used multiple methodologies to measure aerosol emission from the respiratory tract before and during CPAP and high-flow nasal oxygen, in an ultra-clean, laminar flow theatre with near-zero background aerosol and recruited patients with COVID-19 to ensure similar aerosol distributions. We conclude that there is negligible aerosol generation with CPAP, that aerosol emission from HFNO is from the machine and not the patient, coughing emits aerosols consistent with airborne transmission of SARS CoV2 and that healthy volunteers are a reasonable proxy for COVID-19 patients.

*Implications of all the available evidence:* CPAP and HFNO should not be considered high risk aerosol generating procedures, based on our study and that of Gaeckle et al. Recorded aerosol emission from HFNO stems from the machine. Cough remains a significant aerosol risk. PPE guidance should be updated to ensure medical staff are protected with appropriate PPE in situations when patients with suspected or proven COVID-19 are likely to cough.

## Introduction

The WHO describes disease transmission as occurring through physical contact, ‘droplet’ inhalation (larger particles which settle in a reasonably short distance) or as ‘airborne’ (smaller particles which can travel as aerosols on air currents, remaining in the air for longer and distributing over a wide area).^1^ Severe acute respiratory syndrome coronavirus 2 (SARS-CoV-2), the virus that causes coronavirus disease 2019 (COVID-19) can be transmitted via aerosols with aerosol emission being the putative mode of transmission for many super-spreading events.^2,3^ Although the exact size range of aerosol particles responsible for airborne transmission (and the ability of virus to survive in these particles) continues to be debated, it is clear that the dispersion of particles smaller than 5μm is largely determined by the room ventilation (air exchange) rate, thereby posing a potential risk to those even not in close contact, especially in poorly ventilated areas.^4^ Quantifying the concentration of particles of this size range is therefore critical for understanding risk of disease transmission. Recent data indicates that fine aerosol particles <600 nm are unlikely to contribute significantly to infectious events as they constitute so little of the volume fraction of respiratory aerosol, and thus the aerosol mass and viral load, so the likelihood of containing a single virion at normal SARS-CoV-2 viral loads is very low.^5–7^

Traditionally, medical procedures have been deemed as ‘aerosol generating’ when there was perceived to be a risk of increased generation of aerosol from the patients’ mucosa or respiratory tract compared to normal breathing, exposing staff and others in the vicinity to risk from inhalation of aerosolised airborne virus. For interventions defined as aerosol generating procedures (AGPs) by national or international bodies, an extra set of infection control precautions are mandated.^8–10^ These often involve: segregating these patients from others, changing personal protective equipment (PPE) to include FFP3 (or N95) masks that limit aerosol inhalation rather than fluid resistant surgical masks (FRSM), ensuring adequate ventilation, and allowing ‘fallow’ time between procedures to allow aerosol to disperse.

All of these mitigation strategies have significant impact on healthcare capacity, costs and potential harms, it is therefore critical to accurately identify whether these procedures truly do generate aerosol.^11^ Our strategy is to identify whether procedures generate appreciable aerosol and if the aerosol number concentration (the number of particles within a unit volume at a particular particle size) is lower than that generated by a cough, the AGP is likely of low risk. Directly determining exposure to the virus released by AGPs is, in many cases, an intractable measurement, limited by sampling efficiency and determination of infectious viral load as well as aerosol concentration.

Oxygen delivery and respiratory support including CPAP and high flow nasal oxygen (HFNO) are used for the management of hypoxaemic respiratory failure complicating COVID-19 pneumonia and other respiratory conditions. This complex respiratory support is delivered in high care medical wards and intensive therapy units (ITU). CPAP and HFNO are currently deemed AGPs by both the WHO and Public Health England (PHE), although the evidence for these recommendations is sparse.^8,10,12,13^

As these are continuously delivered, current guidance stipulates the need to cohort these patients and universal FFP3 usage in any setting where a patient is receiving CPAP or HFNO. However, universal FFP3 usage is not currently recommended on general medical inpatients with COVID-19 on oxygen via nasal cannula, despite the potential risks from breathing, speaking, and coughing in this setting.

In this study, we set out to quantify aerosol generation in both CPAP and HFNO and compare it to breathing, speaking and coughing without these supports.

## Methods

### Study design

This was performed as part of the wider AERATOR study to assess the risk of aerosolised transmission of SARS-CoV-2 in healthcare settings. Ethical approval was given by the North West Research Ethics Committee (Ref: 20/NW/0393, HRA Approved 18/9/20).

### Aerosol measurement

Aerosol measurements were recorded using two devices simultaneously: an Optical Particle Sizer (OPS) and Aerodynamic Particle Sizer (APS). Technical specifications were detailed in a previous publication from our group but are replicated here.^14^ The key difference was the use of a shorter length of sampling tube (0.45m) from the sampling funnel through to the instrument inlet as patients were seated for this experiment, not supine.

The APS (TSI Incorporated, model 3321, Shoreview, NM, USA) measures aerosol at a sampling flow rate of 1 L min^-1^ with accompanying sheath flow of 4 L min^-1^. The APS reports the aerodynamic size of particles in an aerosol plume, size-resolving aerosol number concentration into 52 size bins ranging from 0.5 µm to 20 µm in diameter with a time integration of 1 s. The size bins are equally spaced in log(diameter) space, apart from the smallest size bin (0.5 - 0.523 µm).

The OPS (TSI Incorporated, model 3330, Shoreview, NM, USA) samples air at 1 L min^−1^ and detects particles by laser optical scattering. The OPS reports the particle number concentration and optical size distribution within the diameter range 300 nm to 10 µm with a time resolution of 1 second. The OPS is widely used for aerosol studies from laboratories / clean rooms to more demanding outdoor environments. It is calibrated by the manufacturer using polystyrene latex spheres and its performance conforms to the ISO standard 21501-4:2018. The reported optical size of the particles is based on an assumed refractive index of pure water at 600 nm wavelength (1.333).

Both the APS and OPS were connected to the same sampling funnel, which was 3D printed (RAISE3D Pro2 Printer, 3DGBIRE, Chorley, UK) from PLA with a maximum diameter of 150 mm, cone height of 90 mm with a 10-mm exit port. Two conductive silicone sampling tubes of 0.3 m length and internal diameter 4.8 mm (3001788, TSI) were connected to the neck of the sampling funnel, with one connected to the APS and the other to the OPS.

For baseline measurements and HFNO, the funnel was placed such that the top of the funnel cone was 1 cm from the participant’s forehead, and the sampling apex of the funnel was 10 cm from the participant’s mouth. For CPAP the sampling apex of the funnel was 10 cm from the exit port or area of greatest facemask leak. Supplement 1 includes some images from the set up to aid visualisation.

### Environmental set up and patient recruitment

Observations were performed in two settings: healthy volunteers were recruited in an ultra-clean laminar flow operating theatre (EXFLOW 32, Howarth Air Technology, Farnworth, UK) with high efficiency particulate air (HEPA) filtration and an air supply rate of 1200 m^3^/s. This ventilation system has a canopy ‘clean zone’ where surgical procedures are performed; the air circulation velocity is 0.2 m.s^−1^ at 1 m above the floor below the canopy and produces 500–650 air changes per hour. All aerosol recordings were performed under the canopy, and the background aerosol concentration was sampled prior to each measurement for a mean sampling duration of 43 s, with mean background number concentrations of 0.00187 (SD 0.00271) cm^-3^ and 0.00330 (SD 0.00395) cm^-3^ reported by the APS and OPS, respectively. Air temperature in theatres was set to 20 °C and humidity between 40 and 60%.

The NIV machine used for delivering CPAP was the Phillips Trilogy 100 and the HFNO machine was the Fisher and Paykel Airvo 2. They were used according to manufacturer instructions. The CPAP circuits used were Armstrong Medical: AMVC1792/032 and Respironics: 1065830, CPAP full face masks were ResMed: AcuCare F1-0 and X, and the nasal cannula used were Fisher and Paykel Optiflow OPT944.

Patients hospitalised with PCR-positive COVID-19 pneumonia were recruited and measurements taken in negative pressure ventilated side-rooms in the infectious disease ward. To reduce the background aerosol number concentration sufficient to allow measurements of baseline procedures in the ward setting, we used a portable HEPA filter (PUREAir R150, PUREAir Limited) with an airflow rate of 1500 m^3^/hour for some patients. This reduced the background concentration for most measurements, although the average background number concentration was 0.351 (SD 0.382) cm^-3^ and 0.407 (SD 0.472) cm^-3^ for the APS and OPS, respectively, still significantly higher than in the laminar flow theatres. Where available, participant height and weight were recorded.

Healthy volunteers were recruited and invited to undergo a protocolised list of procedures in laminar flow environment of an operating theatre (see Supplement S1 for full procedural list). These procedures included: tidal breathing, speaking (with/without mask), coughing (with/without mask), and receiving CPAP (non humidified, full face mask, Continuous Positive Airway Pressure 15cmH_2_0), and HFNO (High flow nasal oxygen). All procedures were performed in the seated position. For some participants, additional measurements were taken (e.g. speaking during HFNO, taking off the CPAP mask). Height and weight were recorded for all volunteers. A sample of the volunteers were invited to have a second measurement on a different date, to ensure replicability. For CPAP, measurements were performed at two areas around the participant: first, at the filtered exit port of the facemask, where most air is exhaled, and then at the point of maximum leak from the mask (measured by an experienced operator). If the maximum leak was less than 50 L/min, a leak was generated by the operator and measurement performed as close as possible to this leak. As with baseline measurements, speaking, breathing, and coughing were recorded during CPAP. Finally, measurements were made as the mask was removed for a small number of participants. CPAP settings were set to our hospital standard (15cm H_2_O pressure), after initial scoping measurements and earlier research found no difference in aerosol emission with changing pressure, oxygen and humidity settings.

For HFNO, measurements were initially performed for both low (30L/min) and high (60L/min) flow rates. As aerosol emission was greater at higher flow rates, subsequent experiments focussed entirely on higher flow rates. As with CPAP, measurements were made during tidal breathing, speaking, and coughing. For some participants, we tested the effect of wearing a surgical facemask over the HFNO nasal cannula on aerosol emission. As described in detail in supplement (Supplement S2), we also performed a set of measurements and experiments with four separate HFNO machines in order to identify the source of aerosol that we recorded during our study.

Hospitalised patients with COVID-19 were recruited by study members and had simple baseline measurements performed (e.g. speaking, breathing, and coughing, both with and without surgical facemasks).

### Statistical analysis

Aerosol generation differs greatly between people, with an approximate log-normal distribution in number concentration.^15,16^ As such, our analysis focussed on comparing the relative aerosol number concentrations from different procedures performed by each individual. We report the number concentration, an intensive property that does not depend on scale (i.e. is independent of the time or volume sampled) as reported by the instruments measured over a sample period, selected to be 1 s. We have reported one of two parameters for each activity: either the peak particle number concentration reported across the full number of samples of the measurement for single, forced exhalations such as coughing (cm-3); or, the mean particle number concentration reported as the average across all samples for continuous activities such as breathing or speaking (cm^-3^). We then visualised size distributions of aerosol emission across the volunteers and compared aerosol emission across activities. Data analysis was performed by collating raw data of sampled aerosol concentration output by the APS and OPS instruments using Aerosol Instrument Manager 9.0 (TSI Incorporated, Shoreview, NM, USA) and Microsoft Excel. A custom-written software in LabVIEW (National Instruments, Texas, USA) was used to automate the analysis process for increased efficiency. For the PCR-positive hospitalised patients with COVID-19, the mean background aerosol number concentration was subtracted from the sampled aerosol number concentration for each activity to account for the non-zero background and to allow comparison with the data from healthy subjects collected under laminar flow. Formal statistical analysis was performed using R 4.0.3 (R foundation for Statistical Computing, Vienna) using t-tests on log-transformed data. As a secondary objective, we wanted to test whether aerosol emission was different for patients with COVID-19 rather than healthy volunteers. For this, we compared activities in hospitalised patients with COVID-19 and in healthy controls. Finally, we tested a sub set of volunteers twice, on different days and with different operators, to assess the intra-person variability in aerosol emission.

## Results

### Overall results and demographics

32 participants were recruited, of which 24 were healthy volunteers, and 8 were hospitalised patients with COVID-19. 13 (57%) of volunteers were female, with a median age of 35 years (IQR 32-40 years), weight of 72kg (IQR 64-79kg), height of 1.74m (IQR 1.64-1.79m) and BMI of 23.6kg/m^2^ (IQR 22.0 −25.5 kg/m^2^).

Hospitalised patients with COVID-19 were older (mean age 55 years, IQR 49-59 years), with 5 men and 3 women. Height and weight were available for 2 patients: both were 170cm tall; one weighed 85kg (BMI: 29.4), the other weighed 139kg (BMI: 48.1kg).

### Volunteer aerosol emission

Table 1 describes the number of times each activity was performed, and on how many volunteers alongside the aerosol emission for each activity. The number of activities does not match on to the number of participants, as some volunteers (n = 6) repeated the assessments on a different day to check repeatability, and some measurements were only performed on certain participants. As described in the methods, the average aerosol number concentration sampled is reported for continuous activities (e.g breathing), whereas the peak number concentration is reported for sporadic activities (cough).

**Table 1:** Aerosol emission produced across all activities in healthy volunteers.

| Oxygen delivery | Activity | Total number of recordings | Total number of volunteers | Aerosol emission (APS, particles/cm<sup>3</sup>)<sup>a</sup> | Aerosol emission (OPS particles/cm<sup>3</sup>)<sup>a</sup> |
| --- | --- | --- | --- | --- | --- |
| Nil | Breathing | 29 | 25 | 0.039 | 0.034 |
| Nil | Speaking | 29 | 25 | 0.113 | 0.094 |
| Nil | Speaking with facemask | 28 | 25 | 0.038 | 0.023 |
| Nil | Cough | 29 | 25 | 1.400 | 1.712 |
| Nil | Cough with facemask | 27 | 22 | 0.075 | 0.207 |
| HFNO | Breathing | 21 | 19 | 2.095 | 2.830 |
| HFNO | Speaking | 21 | 19 | 1.509 | 1.998 |
| HFNO | Cough | 21 | 19 | 3.333 | 4.176 |
| HFNO | Cough with facemask | 10 | 10 | 0.637 | 0.835 |
| CPAP | Breathing sampling at area of greatest natural leak | 21 | 19 | 0.009 | 0.009 |
| CPAP | Breathing sampling at exit port | 21 | 19 | <0.000 | < 0.000 |
| CPAP | Speaking sampling at exit port | 9 | 8 | <0.000 | < 0.000 |
| CPAP | Cough sampling at exit port | 20 | 18 | 0.003 | 0.002 |
| CPAP | Cough sampling at leak | 18 | 16 | 0.029 | 0.011 |
| CPAP | Removing CPAP mask | 6 | 6 | 0.342 | 0.417 |
| a: This is the geometric mean across individuals; average particles/cm<sup>3</sup>/s for continuous activities, peak particles/cm<sup>3</sup> for sporadic activities |  |  |  |  |  |

Correlation between the APS and OPS devices was high (r = 0.98, unlogged, r = 0.80 logged), despite the differing methodologies and flow rates (5 L/min vs 1 L/min), suggesting the rate of flow of air did not strongly influence the results. Further analysis reports the APS figures in the text only, except where stated.

Figures 1a and 1b show the aerosol concentrations of each activity for volunteers, as reported by the APS, (see Figure S3 for the OPS) and show the clear variation in aerosol concentrations (and large variation between people). For baseline measurements, speaking produced more aerosol than breathing, and wearing a fluid resistant surgical masks (FRSM) significantly reduced aerosol emission in both speaking (0.113 vs 0.038, p = 0.002), and coughing (1.40 vs 0.075, p < 0.001). The nature of a cough being a forced, short-lived exhalation of high flow rate makes it challenging to quantify absolute numbers of particles generated using sampling instruments with relatively low flow rates (1 L/min and 5 L/min for the OPS and APS, respectively). However, we can report the peak concentration of aerosol observed in any sample directly after a cough (equivalent to the peak observed in any 1 s of acquisition) and compare this to the mean concentration generated through speaking and breathing. This analysis shows that a single cough generates at least 10-fold more particles at the peak concentration relative to the mean concentration for speaking or breathing (geometric mean concentrations of 1.400 cm^-3^, 0.113 cm^-3^ and 0.039 cm^-3^ for cough, speaking and breathing respectively). The aerosol concentrations as sampled simultaneously by the OPS are reported in analogous box and whisker plots in Figure S3. The mean size distribution of particles sampled by the APS during the baseline measurements is reported in Figure S4.

**Figure 1:**
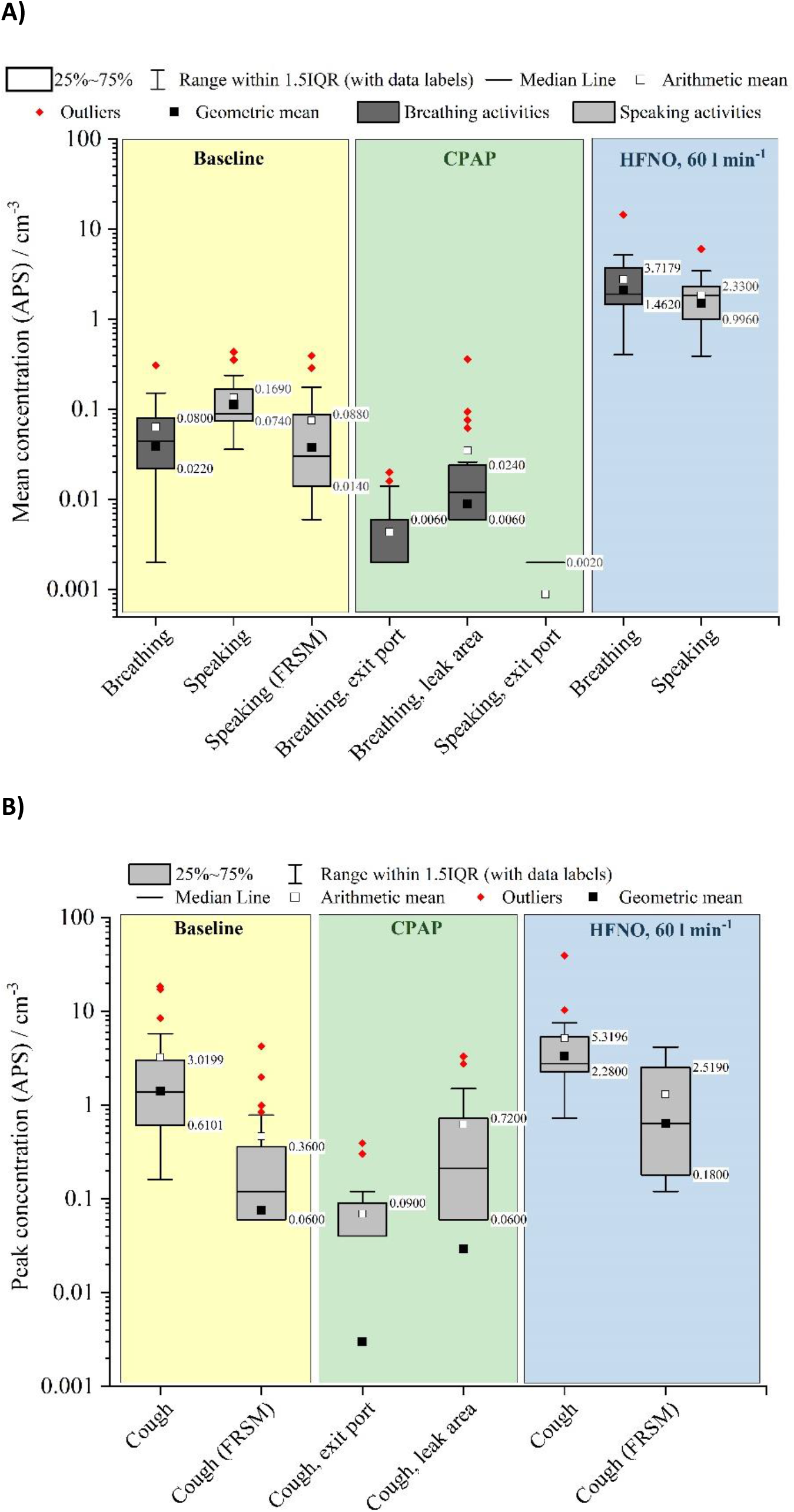
The aerosol number concentration sampled by an APS during baseline activities, CPAP or HFNO, with a) reporting the mean concentration sampled during breathing and speaking and b) reporting the peak concentration sampled during coughs.

### NIV-CPAP

As shown in Figure 1a and 1b, aerosol emission from patients receiving CPAP is greatly reduced compared to all activities. Even with a large induced air leak (>50 L/min), the aerosol emission measured over that leak for coughing was lower than in participants not receiving CPAP 0.029 vs 1.40 particles/cm^3^; p <0.001). At the filtered exit port, the aerosol emission was negligible, and much reduced compared to those not wearing CPAP full face masks (p-values for all comparisons <0.001).

Removal of the mask was associated with some aerosol emission, but this was significantly less than a cough in a healthy volunteer (peak of 0.38 particles /cm^3^ vs 1.57 particles/cm^3^, p <0.001). In summary, CPAP was not associated with increased aerosolisation, but conversely was associated with much lower recorded aerosol number concentrations across all settings.

### HFNO

In contrast to CPAP, HFNO was associated with increased aerosol concentrations compared to baseline measures (p<0.001 for 30 L/min vs baseline, p <0.001 for 60 L/min vs baseline for all comparisons). Higher flow rates (60 L/min) were associated with higher reported aerosol concentrations than lower flow rates (30 L/min) for speaking (0.29 vs 1.71 particles/cm^3^, p <0.001), breathing (2.40 vs 0.33 particles/cm^3^, p <0.001), but not coughing (3.70 vs 2.61 particles/cm^3^, p = 0.155), nor coughing with a surgical facemask (0.73 vs 0.34 particles/cm^3^, p = 0.08).

However, the characteristics of the aerosol emissions during HFNO was not consistent with production of aerosol from respiratory tract or mucosal surfaces. We performed a set of experiments to assess the source of this aerosol, with full experimental detail in a supplementary appendix (See “Aerosol Concentrations Generated by HFNO” and Fig. S5, S6 and S7). Briefly, the aerosol concentration produced by HFNO varied significantly between four different machines tested, with high concentrations of aerosol emission being recorded in two of the four machines despite not being attached to the patient. A zero particle filter attached between the nasal cannulae and the machine reduced this aerosol emission. Secondly, the size distribution of particles sampled when patients performed breathing and speaking whilst wearing HFNO clearly represent the additive effect of two separate size distributions of particle populations: one from the respiratory tract and one arising from the HFNO device (Fig. S8). Given the size distributions of particles generated solely by the HFNO device is lognormal with the peak in the sub-micrometre diameter range, these particles are expected to follow the HFNO airflow through the nasal cavity and are exhaled without impaction in the upper airways. Thus, whilst there are nominally more aerosol particles sampled whilst a patient breathes or speaks during HFNO than without the oxygen delivery, we do not envisage that the additional submicron particles generated by the HFNO would pick up any mucosal material from the participant to pose an infection risk of COVID-19. This conclusion is supported by the similarity in the sub-micrometre size distributions from the HFNO directly and then with the patient. In summary, although some HFNO machines do produce a significant amount of aerosol, this aerosol is generated by the machines and is not of clinical relevance and does not pose a risk of infection.

### COVID patients vs healthy volunteers

In total, 8 patients were recruited with COVID-19. Measurement of aerosol concentrations generated by these patients was technically difficult due to infection control requirements, and the air quality of the room, necessitating HEPA filtration for some participants. 4 participants were on standard, low flow, nasal oxygen, while the others were breathing room air. For 4 participants, the background aerosol concentration in the room was higher than median concentrations generated by speaking and breathing measured under laminar flow, preventing any assessment of concentrations from speaking and breathing by COVID-19 positive patients.

Figure 2 shows the difference in aerosol emission for each activity between COVID-19 patients and healthy participants, with Table 2 showing the average emission for each group and activity. Broadly speaking, hospitalised patients COVID had higher aerosol emission than healthy volunteers. This higher emission from COVID patients is due in part to a reporting bias towards higher-emitting patients, who produced aerosol concentrations observable above the background.

**Figure 2:**
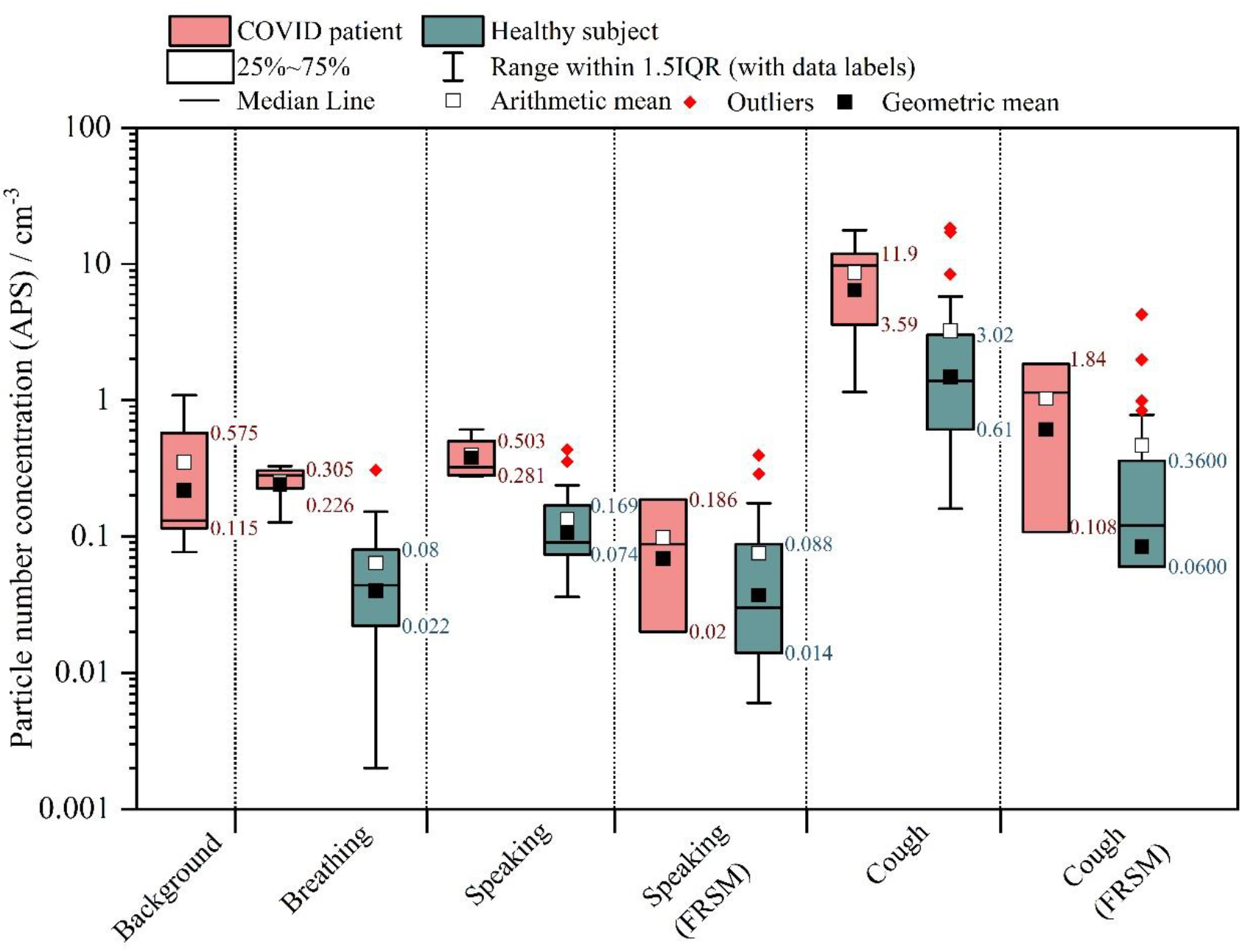
Box and whisker plot comparing the aerosol sampled by an APS when baseline activities are performed by healthy subjects and by PCR-positive patients with COVID-19.

Importantly, the size distribution of aerosol particles in patients with COVID-19 was very similar to healthy volunteers (Figure S3). Breathing, speaking and coughing all generated aerosol particles in a lognormal size distribution with the peak in the 0.5 – 1 µm diameter range, consistent with previous reports of the size distribution of respiratory aerosol emissions.^14,16–18^ This supports the use of healthy volunteers as proxies for patients infected with SARS-CoV-2

### Repeated measurements

For a subset of healthy volunteers (n = 6), repeated measurements were made. These were all performed on a different day, more than 1 month after the first measurement. In total, there were 116 measurements repeated, 76 with the APS and 40 with the OPS. In general, correlation with the original measurement was high (r = 0.71 on logged data), although this was driven by strong correlation in breathing (r = 0.81), rather than speaking (r = 0.17 on logged data) and coughing (r = 0.38 on logged data) suggesting aerosol concentrations from breathing are relatively consistent for any individual recorded over a period of time. Figure S1 and S2 show this data, coloured by participant (S1) and activity (S2).

## Discussion

### Summary

This study comprehensively assessed the aerosol generation from NIV (continuous positive airways pressure) and high flow nasal oxygen (HFNO), as compared to normal breathing, speaking and coughing. CPAP was clearly associated with reductions in aerosol number concentrations, even with large leaks around the face mask (>50L). The exit filter eliminated nearly all aerosol emission during coughing and breathing, suggesting well fitted CPAP is associated with negligible aerosol generation. For HFNO, aerosol concentrations were higher than baseline recordings. However, this additional aerosol is generated from the device, and not from the patient, and is unlikely to be of clinical relevance. These oxygen delivery systems should not be deemed AGPs, and provide no greater (perhaps less, with CPAP) risk to health care staff relative to patients breathing, coughing, and talking without support.

This is the first study to report on aerosol emission from patients with active COVID-19, with previous work on primates only.^19^ While the data available for analysis suggests that peak aerosol concentrations from coughs are higher than recorded from healthy volunteers without COVID-19 who were recruited in a laminar flow theatre, the background aerosol concentration on the ward was too high to make reliable inference about breathing and speaking. Some patients had lower emissions, which were not detectable above the non-zero background. However, the background level enabled estimations for maximum possible aerosol emission which were not that different from healthy volunteers.

We also report the results of repeated measurements with healthy volunteers. These were all performed more than one month after the initial measurement and show high correlation (r = 0.71) with the initial measurements, although this was driven by strong correlations in breathing results as opposed to speaking and coughing.

In summary, our data (in concert with prior research on AGPs,^12,14^ epidemiological data suggesting lower risk to ITU staff,^20–24^ and with viral loads higher earlier in infection, when patients are more often on the general wards^25^), strongly support an equal or even greater risk of SARS-CoV-2 infection via the aerosol route for health care workers providing care to patients with COVID-19 on acute medical units, general medical wards or in the emergency department rather than those on critical care wards, suggesting policy on PPE in particular should be re-evaluated.

### Strengths and weaknesses

#### This study has multiple strengths

Firstly, it was performed in ultra-clean laminar flow theatres, with very low background, allowing for accurate quantification of aerosol emission. Secondly, the strong correlation between both aerosol measurement modalities provides confidence that the aerosol measurements are reliable. Thirdly, repeated measurements and the recruitment of patients with active COVID-19 allows for greater interpretation of clinical implications. Finally, the protocol reflects usual clinical care and is directly translatable to the health service delivery.

Common to all volunteer studies, much of the data comes from healthy participants. However, this was supplemented with data from patients with COVID-19; and while aerosol generation was increased (within the limitations of the background concentration), the particle size distributions were very similar, supporting the principle that aerosol emission is not different in nature between patients and healthy volunteers. Secondly, the measurement methodology employed by the APS and OPS uses relatively low flow rates (1 L/min and 5 L/min, respectively). These mean that very short, high impact aerosol emissions (e.g. cough) may be hard to quantify. However, this applies to all aerosol measurements using APS and OPS technology, and does not limit relative comparison between oxygen delivery systems. Finally, while we tested CPAP at one time point and one single setting, this reflects current clinical practice and, initial experimentation (unpublished), and previous work by Gaeckle et al. found no difference between different CPAP pressures. Finally, our CPAP NIV system uses full face masks with exhalation filters, which are standard care in NHS hospitals. We cannot extrapolate to CPAP face masks without exhalation filters, although aerosol concentrations recorded from the leak (unfiltered), was reduced compared to coughing without CPAP.

It is important to note that some of the reduction of aerosol emission in breathing and speaking recorded during leak from the CPAP mask may be driven by a dilution effect (i.e it may be that a similar number of particles are produced in CPAP, but the additional volume of air provided by the CPAP machine dilutes this leading to a reduction in measured aerosol emission). However, such dilution is equivalent to also increased air exchange rate in a room and will effectively reduce the associated risks of airborne transmission.

We have chosen not to correct the reported particle concentrations sampled during each procedure to account for the effect of dilution by the airflow, because the relative flow rate between each subject’s different exhalation events compared to CPAP and HFNO are ill-defined. Thus, the uncorrected aerosol number concentrations as sampled by the APS and OPS do not represent the absolute quantity of particles generated by each activity, but can be used as a measure of the risk to a healthcare worker in the vicinity of the activity.

As activities such as coughing are very forceful and short lived, these were analysed separately to the continuous activities (e.g. breathing): short transient activities are observed as a rapid rise in the reported number concentration, but decay away over a few sample measurements (typically equivalent to 10-15 s for a cough) as the aerosol dissipates from the sampling funnel and is diluted by the clean room air. While reporting the intensive property of concentration allows us to compare relative yields from aerosol generating procedures, it is important to note that estimating absolute yields or fluxes (extensive properties) requires knowledge of the volumetric flow rates for the gas in which the aerosol is dispersed. These present an additional challenge to measure. Although it is possible to report the absolute number of particles counted by the instruments (given we know their sampling volumetric flow rates), we cannot conclude that this is equivalent to the total aerosol yield without knowing the volumetric flow rate at aerosol source.

### Comparisons with previous literature

There are few previous published studies of aerosol generation from oxygen delivery systems and respiratory support. Hui and colleagues performed experiments on CPAP, HFNO and simple nasal cannulae using smoke inside a human patient simulator and measured the distance and trajectory of the smoke. ^13,26^ As commented by others, ^12^ unsurprisingly the higher pressure forced the smoke longer distances, through small ports in masks.

The most similar, recent study was performed by Gaeckle et al.^12^ In this study, measurements of the aerodynamic number concentration (by APS) were reported from ten healthy volunteers in a negative pressure room (∼15 air exchanges per hour), with an additional portable HEPA filter added to reduce background aerosol concentration. A similar protocol was used to ours, although they measured simple nasal cannulae and changes in respiration. Importantly, they reported a background aerosol concentration of ∼0.060 particles/cm^3^ (compared to zero under laminar flow), larger than we report for many activities (including breathing and speaking with a facemask). As well as a high background, the aerosol number concentration was highly variable in their study (see Figure 4 and E3 from reference 11, and figure 3 here for comparison). This variability makes reporting accurate aerosol concentrations for short events (e.g. a cough) challenging, as we noted in recruiting our COVID-19 patients.

**Figure 3:**
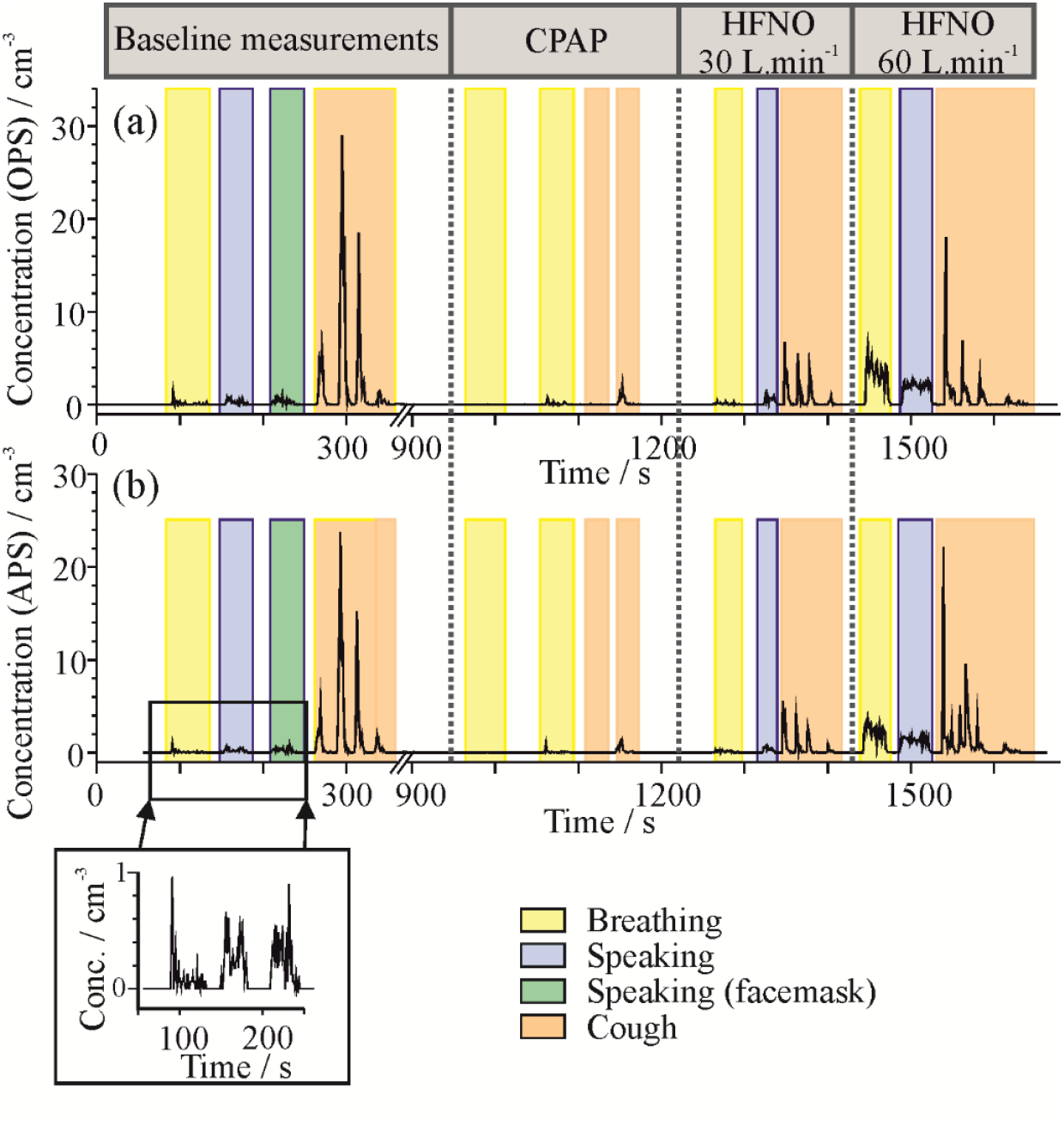
Example of the time series of OPS (a) and APS (b) number concentrations sampled during a measurement of one healthy subject performing baseline activities, followed by CPAP then HFNO.

Consistent with our study, Gaeckle et al. reported NIV to be non-aerosol generating. However, by contrast they did not identify increased aerosol emission with HFNO. In two of the machines we tested, no aerosol emission was identified, whereas the other machines produced significant (but different) aerosol concentrations. This, alongside our experiments detailed in Supplement S2 show that aerosol is generated from certain HFNO machines, and the receipt of HFNO doses not increase aerosol emission from the respiratory tract. This also suggests there is significant variation in aerosol emission between HFNO machines, accounting for the differences between our and their work.

Emerging evidence supports the risk of COVID-19 infection to be lower among critical care staff (who generally manage patients on both CPAP and HFNO) than other healthcare staff (OR 0.29; 95%CI 0.13-0.57 from our hospital^20^, similar results from other cohorts^19,21–24^). There are many plausible explanations (fewer patients per clinician, patients presenting later in disease course), but one of the most likely is the use of more highly protective FFP3 masks by critical care staff.

The analysis presented in this paper, along with other work from our group identifying that intubation does not also generate significant aerosol^14^ and this epidemiological data, suggest that the current strategy of supporting critical care staff but not general ward staff with FFP3 masks to mitigate the risks associated with AGPs is misplaced. Risk of aerosolization from patients with COVID-19 is at least as likely in medical wards than in intensive care units. Indeed, it may actually be much more likely given the protective effect of closed circuit ventilation and the likely protective effect of CPAP masks we have identified. Finally, the viral load dynamics suggest the highest infectious viral load is at or around the time of symptoms, and falls off after.^25^ As patients present to critical care units later, this provides further support for increasing risk for general ward and admissions staff.

### Implications for clinical practice and policy

This study strongly supports re-evaluation of guidance listing CPAP as a high risk AGP, especially when considered alongside recent work from others. PPE policy around masks should be guided by exposure to potentially infectious aerosol, and not be guided by non-invasive ventilation strategies. In the COVID-19 pandemic, acute and general medical ward staff managing Covid-19 patients are just as likely as intensive care staff to be exposed to infectious aerosol, and should be protected accordingly.

## Conclusions

NIV such as CPAP via a filtered mask actually reduces aerosol emission compared to normal breathing. HFNO does generate additional aerosol, however this aerosol is generated from the machine and not the patient, and is unlikely to pose extra clinical risk given the size (<1um). Cough appears to generate significant aerosols in a size range compatible with airborne transmission of SARS-CoV-2. Policy around personal and protective equipment should be updated to reflect these adjusted risks.

## Data Availability

Raw data from the APS and OPS is available on reasonable request from the corresponding author.

