## Supplement S1 for "Aerosol emission from the respiratory tract: an analysis of relative risks from oxygen delivery systems"

### SUPPLEMENTARY APPENDIX 1

Hamilton F*[1,2], Gregson F*[3], Arnold D[4], Sheikh S[3], Ward K[5], Brown J[6], Moran E[1],White C[7] AERATOR group, Bzdek B[3], Reid J[3], Maskell N[4#], Dodd JW[4#]

*These authors contributed equally

#These authors contributed equally

1. Infection Sciences, North Bristol NHS Trust, BS10 5NB
2. MRC Integrative Epidemiology Unit, University of Bristol, BS8 2HW
3. Bristol Aerosol Research Centre, School of Chemistry, University of Bristol, BS8 1TS
4. Academic Respiratory Unit, University of Bristol, BS10 5NB
5. Physiotherapy Department, North Bristol NHS Trust, BS10 5NB
6. Intensive Care Unit, North Bristol NHS Trust, BS10 5NB
7. Research and Innovation, North Bristol NHS Trust, BS10 5NB

AERATOR group (in alphabetical order): The AERATOR group consists of (in alphabetical order): Arnold, D; Brown, J; Bzdek, B; Davidson, A; Dodd, JW; Gormley M; Gregson, F; Hamilton, F; Maskell, N; Murray, J; Keller, J; Pickering, A.E; Reid, J; Sheikh, S; Shrimpton, A.

Corresponding author:

James Dodd

Supplementary Appendix 1:

List of procedure, plan and set up:

| Aerosol instruments on (baseline 30 s) | Measurement position |
|---|---|
| Breathe into funnel (30 s) | Funnel 10cm from mouth |
| STOP step back from funnel | |
| Speak into funnel (30 s) ~ 75 dB | Funnel 10cm from mouth |
| STOP step back from funnel | |
| Speak (30 s) **WITH FRSM** ~ 75 dB | Funnel 10cm from mouth |
| STOP step back from funnel | |
| Cough, then step back for 20 seconds | Funnel 10cm from mouth |
| Cough, then step back for 20 seconds | Funnel 10cm from mouth |
| Cough, then step back for 20 seconds | Funnel 10cm from mouth |
| Cough, **WITH FRSM**, then step back for 20 seconds | Funnel 10cm from mouth |
| STOP step back from funnel + place CPAP mask on | |
| Manouvere so CPAP exit port in funnel (30s) +<br>breathing | CPAP exhalation port in funnel |
| STOP step back from funnel | |
| Manouvere so CPAP area of greatest leak in funnel (30s) + breathing | Funnel 10cm from area of greatest leak |
| STOP step back from funnel | |
| Manouvere so CPAP exit port in funnel (30s) +<br>speaking | CPAP exhalation port in funnel |
| STOP step back from funnel | |

| | |
|---|---|
| Manouvere so CPAP area of greatest leak in funnel (30s) + speaking | Funnel 10cm from area of greatest leak |
| STOP step back from funnel | |
| Manouver so CPAP exit port in funnel, cough, then stand back 20 seconds | CPAP exhalation port in funnel |
| STOP step back from funnel | |
| Manouver so CPAP exit port in funnel, cough, then stand back 20 seconds | CPAP exhalation port in funnel |
| STOP step back from funnel | |
| Manouvere so CPAP area of greatest leak in funnel, cough, then stand back 20 seconds (30s) | Funnel 10cm from area of greatest leak |
| STOP step back from funnel | |
| Manouver so CPAP area of greatest leak in funnel, cough, then stand back 20 seconds | Funnel 10cm from area of greatest leak |
| STOP step back from funnel - remove CPAP mask | |
| Place NHF02 on at low flow with entrained air - measure tidal breathing (30s) | Funnel 10cm from mouth |
| STOP step back from funnel | |
| Place NHF02 on at low flow (20L) with entrained air - measure tidal breathing (30s) | Funnel 10cm from mouth |
| STOP step back from funnel | |
| Place NHF02 on at high flow (60L) with entrained air - measure tidal breathing (30s) | Funnel 10cm from mouth |
| STOP step back from funnel | |
| Place NHF02 on at high flow (60L) with entrained air - speaking (30s) | Funnel 10cm from mouth |

| | |
|---|---|
| STOP step back from funnel | |
| Place NHF02 on at high flow (60L) with entrained air - speaking (30s) + FRSM | Funnel 10cm from mouth |
| STOP step back from funnel | |
| Cough, then step back for 20 seconds (on 60L/min) | Funnel 10cm from mouth |
| Cough, then step back for 20 seconds (on 60L/min) | Funnel 10cm from mouth |
| Cough, then step back for 20 seconds (on 60L/min) | Funnel 10cm from mouth |
| STOP step back from funnel | |

Note 1: Because coughs tend to decrease in strength, for some participants, we did a cough wearing a FRSM first in some participants.

Note 2: Speaking was asking patients to count from 1-100, at a set cadence and aiming for a similar volume between participants.

Photos of the set up:

1. An example of where the volunteer would sit in relation to the measurement devices

PICTURE REMOVED DUE TO MEDRXIV POLICY – PLEASE AWAIT FULL PAPER OR CONTACT CORRESPONDING AUTHOR FOR IMAGE

2. A second image of the set up

PICTURE REMOVED DUE TO MEDRXIV POLICY – PLEASE AWAIT FULL PAPER OR CONTACT CORRESPONDING AUTHOR FOR IMAGE

3. A healthy volunteer receiving HFNO

PICTURE REMOVED DUE TO MEDRXIV POLICY – PLEASE AWAIT FULL PAPER OR CONTACT CORRESPONDING AUTHOR FOR IMAGE
