## Supplement S2 for "Aerosol emission from the respiratory tract: an analysis of relative risks from oxygen delivery systems"

### SUPPLEMENTARY APPENDIX 2

Hamilton F*[1,2], Gregson F*[3], Arnold D[4], Sheikh S[3], Ward K[5], Brown J[6], Moran E[1],White C[7] AERATOR group, Bzdek B[3], Reid J[3], Maskell N[4#], Dodd JW[4#]

*These authors contributed equally

#These authors contributed equally

1. Infection Sciences, North Bristol NHS Trust, BS10 5NB
2. MRC Integrative Epidemiology Unit, University of Bristol, BS8 2HW
3. Bristol Aerosol Research Centre, School of Chemistry, University of Bristol, BS8 1TS
4. Academic Respiratory Unit, University of Bristol, BS10 5NB
5. Physiotherapy Department, North Bristol NHS Trust, BS10 5NB
6. Intensive Care Unit, North Bristol NHS Trust, BS10 5NB
7. Research and Innovation, North Bristol NHS Trust, BS10 5NB

AERATOR group (in alphabetical order): The AERATOR group consists of (in alphabetical order): Arnold, D; Brown, J; Bzdek, B; Davidson, A; Dodd, JW; Gormley M; Gregson, F; Hamilton, F; Maskell, N; Murray, J; Keller, J; Pickering, A.E; Reid, J; Sheikh, S; Shrimpton, A.

Corresponding author:

James Dodd

### <u>Repeat Measurement Data</u>

A subgroup of the healthy subjects (*n*=6) were invited to repeat the baseline measurements one month later. The comparison between aerosol concentrations sampled during the original measurement and later measurement are shown in Figure S1 and S2, showing the correlation between the two measurements (r = 0.71 on logged data). This suggests that the aerosol concentrations generated during healthy respiratory emissions are relatively consistent for any individual over a period of time.

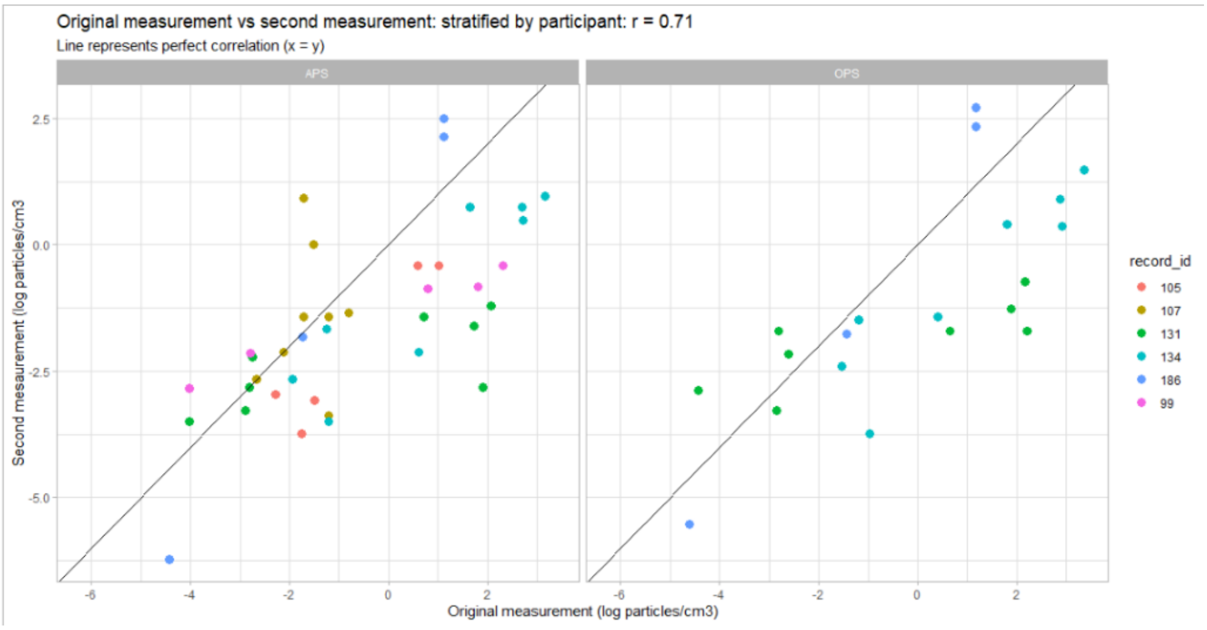


**Figure S1:** The aerosol number concentration sampled by an APS during baseline measurements for six subjects compared to that for a repeat measurement at a later date. For breathing and speaking activities, the mean concentration per sample (equivalent to 1 s acquisition) is reported; for coughing the peak concentration sampled during any sample (1 s acquisition) is reported. Data are classified by colour according to the subject (displayed in legend).

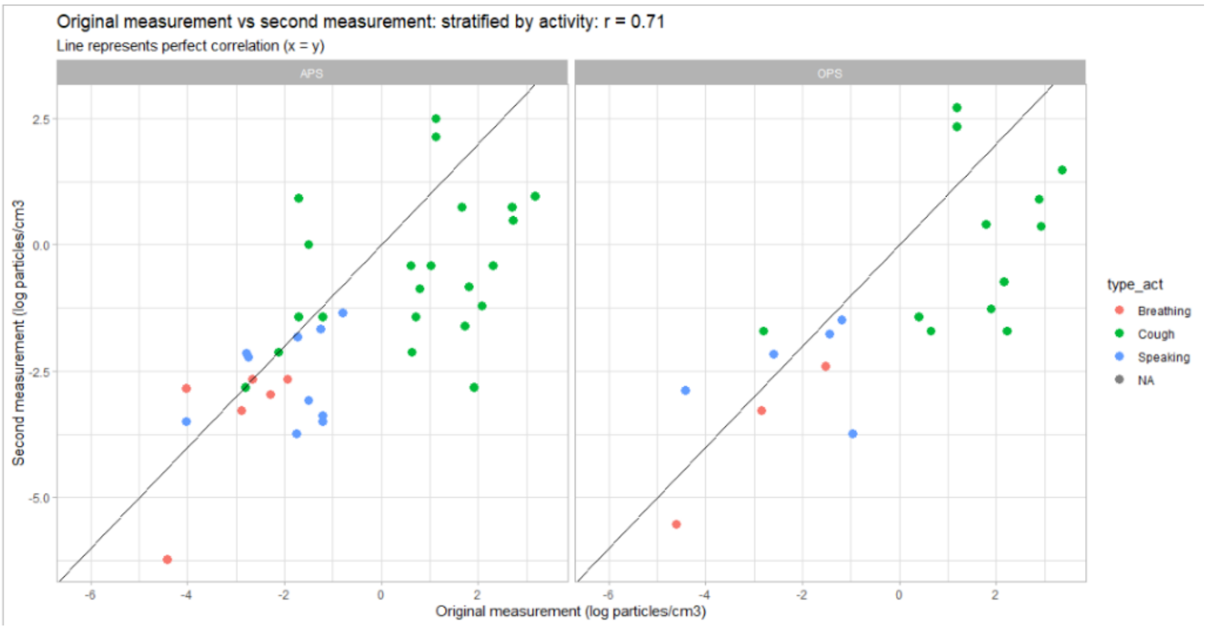


**Figure S2:** The aerosol number concentration sampled by an APS during baseline measurements for six subjects compared to that for a repeat measurement at a later date. For breathing and speaking activities, the mean concentration per sample (equivalent to 1 s acquisition) is reported; for coughing the peak concentration observed during any sample (1 s acquisition) is reported. Data is classified by colour according to the type of respiratory emission (displayed in legend).

### <u>OPS Measurement Data</u>

Aerosol concentrations were recorded simultaneously with an OPS and APS. The box and whisker plots for the OPS measurements (equivalent to Figure 1) are shown in Figure S3.

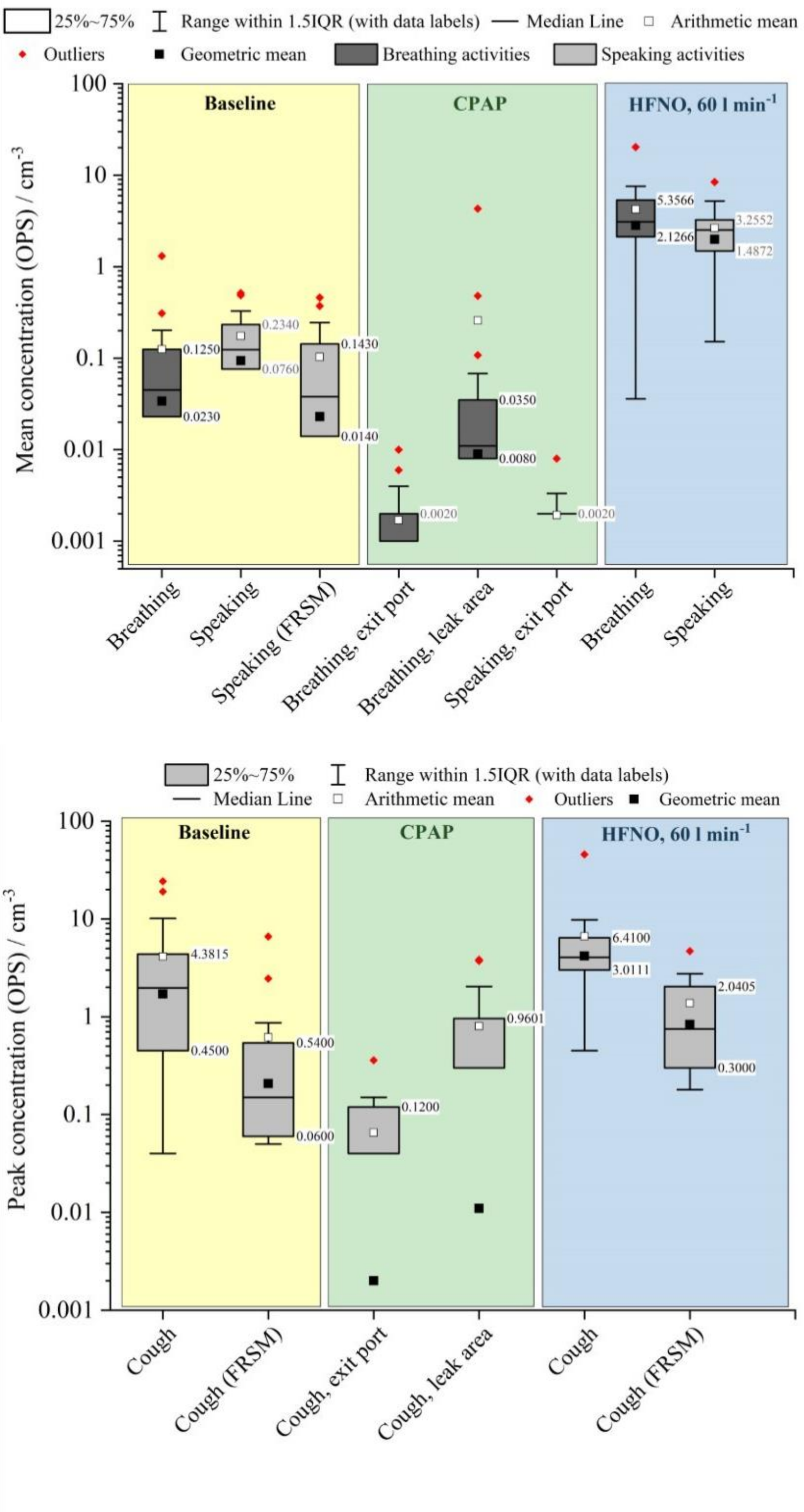


**Figure S3:** The aerosol number concentration sampled by an OPS during baseline activities, CPAP or HFNO, with a) reporting the mean concentration sampled during breathing and speaking and b) reporting the peak concentration sampled during coughs.

### Size Distributions of Sampled Aerosol

The mean size distributions sampled by the APS from all subjects during different measurements are shown in Figure S4. The respiratory aerosol particles form a lognormal size distribution, with the majority of particles generated in the sub-micrometre diameter range for coughing, breathing and speaking. The aerosol particles sampled when a subject spoke or coughed while wearing a fluid-resistant surgical facemask had a reduced number concentration compared to when wearing no mask (0.113 vs 0.038, p = 0.002 and 1.40 vs 0.075, p < 0.001). This is particularly evident when looking at the size distributions in the diameter range 0.5 - 1.5 µm (Fig. S4a). The particles generated by a cough during CPAP, sampled at the area of greatest leak (orange squares) appear to show a similar size distribution to a regular baseline cough (red squares), but are reduced in number concentration.

The mean size distribution of particles generated by healthy patients breathing, speaking and coughing is compared to that from patients infected with COVID-19 in Fig. S4b. The sample size of the COVID-19 patients is reduced (*n* for each activity reported in Table 2) and only shows data for which the aerosol peak could be observed above the high background aerosol concentration, so the number concentration appears greater than that for healthy subjects. However, the size distribution of the sampled particles is similar, with the peak of the lognormal distributions for each activity aligning at a comparable aerodynamic diameter for healthy subjects vs. COVID-19 subjects.

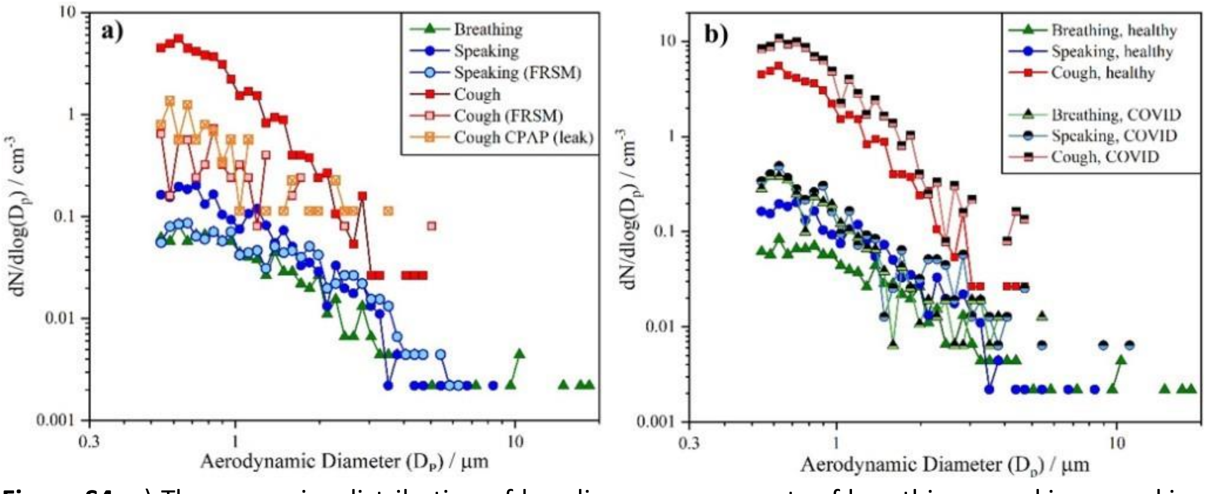


**Figure S4:** a) The mean size distribution of baseline measurements of breathing, speaking, speaking with a face mask, coughing, coughing with a facemask (all *n*=25) and coughing during CPAP, sampling at the area of greatest leak (*n* = 17) b) The mean size distribution of baseline measurements performed by healthy subjects compared to by PCR-positive patients with COVID-19.

### Aerosol Concentrations Generated by HFNO

Considering the high aerosol level for all baseline activities conducted with 60 L min$^{-1}$ HFNO compared to CPAP or no oxygen delivery system (Figure 1), the aerosol source from HFNO, both with and without a subject present, was investigated. One subject generated a mean aerosol concentration (sampled by an APS) during baseline speaking of 0.162 $cm^{-3}$, which increased to 1.7 $cm^{-3}$ when speaking during HFNO (Figure S5). Given that the mean concentration sampled when the HFNO cannula with 60 L $min^{-1}$ humidified air (no subject present) was held 10 cm from the sampling funnel apex was 1.312 $cm^{-3}$ (Fig.S2), it appears that the additional particles sampled during the subject's measurement of speaking during HFNO have originated from the HFNO device and were sampled in addition to the respiratory aerosol from speaking. Likewise, the mean baseline breathing aerosol concentration for this subject was 0.002 $cm^{-3}$, which rose to 2.4 $cm^{-3}$ when the patient breathed during HFNO.

Introducing a 0-particle HEPA capsule filter (removal rating: 1.2 mm Versapor® membrane filteration area: 860 $cm^2$ , set-up shown in Figure S6) in series with HFNO tubing between the humidifier and the nasal cannula reduced the concentration of sampled aerosol from the HFNO device alone (no subject present) by two orders of magnitude to 0.005 $cm^{-3}$. When the subject was re-introduced to undergo HFNO, with the HEPA filter still present in series, the mean particle number concentration sampled was 0.006 $cm^{-3}$ during the subject's breathing, three orders of magnitude lower than that with no HEPA filter inserted in the HFNO tubing. Therefore, it can be concluded that a large proportion of particles detected from HFNO activities are generated by the HFNO air flow rather than the subject. These trends were also observed in the sampled OPS concentration data (not shown).

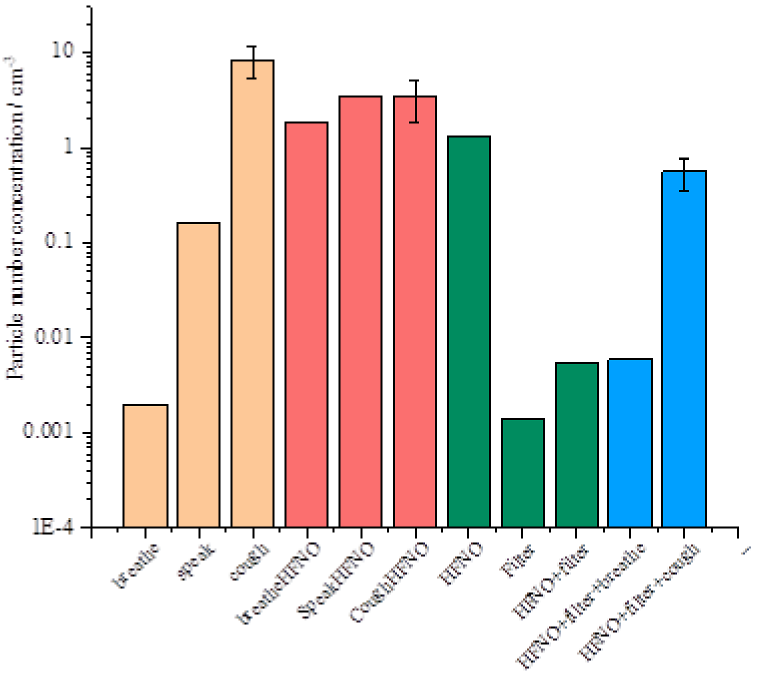


**Figure S5:** APS total particle count detected from 1 second of activity / cm-3 for breathing and speaking and peak height cough data / cm-3 from one subject assessing particle source during HFNO delivery. Baseline measurements (cream) show breathing and speaking and coughing data without HFNO. Baseline measurements whilst receiving HFNO delivery (coral) for subject are greater. Addition of the 0-particle HEPA filter without the subject present measurements are shown in green. The subject was then introduced with the filter in series (blue). The sustained, timed activities (breathing, speaking, HFNO device alone) have the mean concentration per sample reported (equivalent to 1 s acquisition)

and concentrations produced during a cough are reported as the peak concentration observed during any one sample (1 s acquisition). Error shows SD.

PICTURE REMOVED DUE TO MEDRXIV POLICY – PLEASE AWAIT FULL PAPER OR CONTACT CORRESPONDING AUTHOR FOR IMAGE

**Figure S6**: Experimental set up showing the insertion of the HEPA capsule filter into the tubing between the HFNO cannula and the HFNO device, sampled by the APS and OPS via the funnel.

In order to further investigate the particle source from HFNO, a subject was invited to repeat the baseline measurements during HFNO but both with and without the HFNO humidifier in operation (Figure S7). There was no appreciable difference between the total particle concentration generated during this subject's baseline measurements during HFNO with the humidifier in operation or during dry HFNO. Speaking whilst receiving humidified HFNO generated a mean concentration of 0.844 cm-3, compared to 0.872 $cm^{-3}$ during dry HFNO. Whilst this additional experiment was performed for only one subject, the results indicate that the HFNO humidifier was not the major source of the particles generated from the HFNO delivery system.

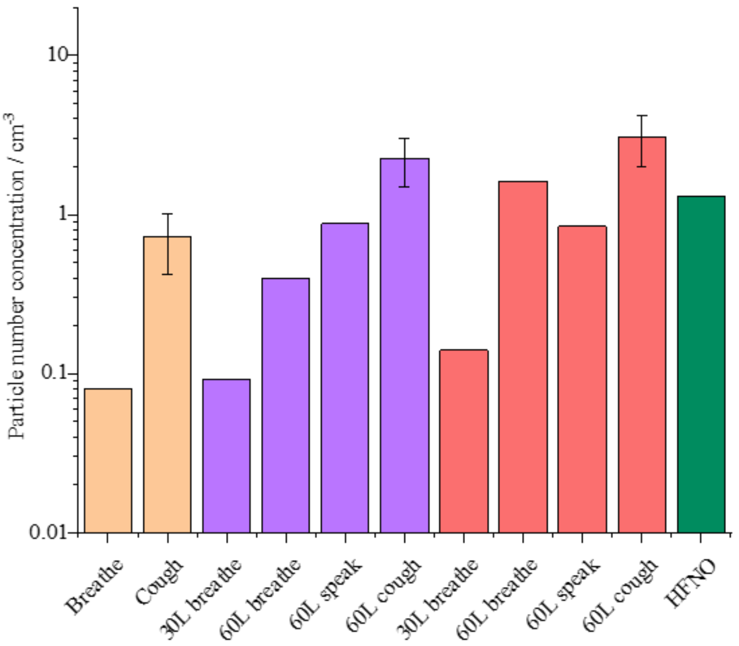


**Figure S7:** The mean concentration sampled by an APS during a subject's baseline measurements (cream), compared to that during dried HFNO (purple) and humidified HFNO (coral). The concentration of particles generated by the HFNO device alone (no subject present) is shown in green. The sustained, timed activities (breathing and speaking) have the mean concentration per sample reported (equivalent to 1 s acquisition) and concentrations produced during a cough are reported as the peak concentration observed during any one sample (1 s acquisition). Error shows SD.

**Size Distributions of Aerosol Concentrations Generated by HFNO**

Investigating the size distribution of particles sampled during subjects performing HFNO can also allude to their origin. Particles generated by the HFNO device alone and sampled by the OPS and APS are shown in Figure S8a (black squares and grey diamonds, respectively). When the subjects (*n*=25) undergoing HFNO breathed into the sampling funnel, the size distribution of particles sampled by the APS (green squares) comprises both those particles sampled directly from the HFNO device (HFNO device #1) as well as those sampled during baseline breathing with no oxygen delivery (dark green triangles). Essentially, during HFNO breathing there appears to be an additive effect of the two populations of particles, both respiratory aerosol and aerosol generated by the HFNO device. The latter, whilst contributing to the nominal increase in particles sampled during HFNO breathing compared to baseline breathing, are not expected to carry COVID-19 transmission risk. As the HFNO device particles are very small in diameter (the peak of the size distribution is well below 0.5 µm and is near zero around 1 µm) these particles are expected to only follow the airflow streamlines through the nasal cavity and be immediately exhaled into the sampled funnel without impaction in the nasal canal or respiratory tract and thus cannot pick up any viral load during transport.

For a subset of the cohort (*n*=6), an additional measurement of breathing during HFNO was performed but with a different HFNO machine (HFNO device #4). The concentration and size distribution of particles generated by this HFNO device alone (no subject present) is different to the first HFNO device, and is shown in the inset to Fig. S8a. When the six subjects undergoing HFNO with the second device breathed into the funnel, there is clearly an additive effect of the two size distributions arising from the HFNO and the breathing.

This additive effect of sampling both the HFNO-generated particles and respiratory particles can be observed for speaking, with both the original HFNO device (*n*=25, Figure S8a) and with the second HFNO device (*n*=6, Figure S8a inset). This serves as evidence that whilst a greater particle concentration is sampled during HFNO compared to baseline measurement with no oxygen support, these additional particles arise from the HFNO device itself and do not carry an associated COVID-19 transmission risk.

The quantity and size distribution of particles generated by the HFNO device is variable between different machines. Figure S8d shows the OPS size distributions of particles originating from four HFNO devices. OPS size distributions are preferentially shown here because the particles are of small diameter that would not be detected by the APS. “HFNO 1”, used in the majority of the HFNO subject data (*n*=23, principal figures in Fig.S8), generated a mean OPS particle concentration of 1.622 $cm^{-3}$, “HFNO 4”, used in the additional HFNO cohort (*n*=6, in the insets of Fig. S8) generated a mean concentration of 3.45 $cm^{-3}$. The cleaner machines “HFNO 2” and “HFNO 3” generated fewer particles, 0.0070 $cm^{-3}$ and 0.020 $cm^{-3}$, respectively, however these devices were not used for a cohort of subjects undergoing HFNO.

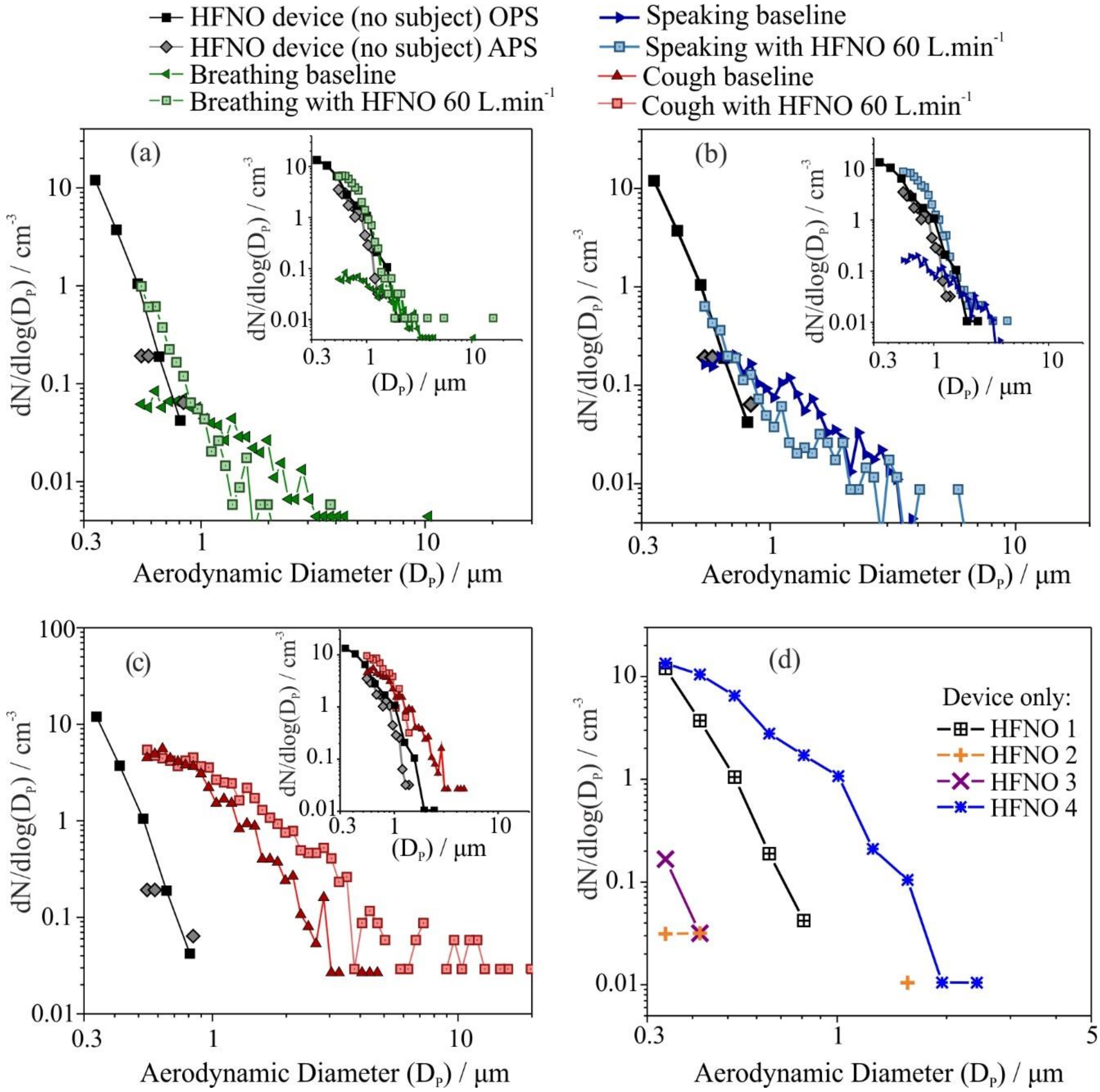


**Figure S8:** The mean size distribution of healthy subjects performing a baseline respiratory activity (*n*=25) and also during 60 L.min$^{-1}$ HFNO (*n*=23), compared to the size distribution of particles generated by the HFNO device and cannula. Inset shows the mean distribution of healthy subjects performing the same baseline activity (*n*=6), but with an alternative HFNO device that generates more particles in a different size distribution. Respiratory activities are breathing, speaking and coughing for a), b) and c), respectively. d) OPS size distributions of particles originating just from the HFNO device for four different devices, where "HFNO 1" is that used for the majority of HFNO datasets (*n*=23) and "HFNO 4" is that used for the additional six subjects in the figure insets. HFNO devices 2 and 3 generate fewer particles but were not used for subject datasets.

The size distribution of particles sampled from a cough during HFNO do not show quite the same additive effect as those for breathing and speaking (Figure S8c and inset). The reasons behind this are unclear. Given that coughs are transient and short lived, different to the sustained respiratory activities of breathing and speaking, there are measurement challenges associated with quantifying the generated concentrations and so the trends of how the size distribution changes for a cough during HFNO may be more complex.
